# The causal effect of serum vitamin D concentration on COVID-19 susceptibility, severity and hospitalization traits: a Mendelian randomization study

**DOI:** 10.1101/2021.03.08.21252901

**Authors:** Zhiyong Cui, Yun Tian

## Abstract

**Background:** Evidence supporting the role of vitamin D in the coronavirus disease 2019 (COVID-19) pandemic remains controversial.

**Methods:** We performed a two-sample Mendelian randomization (MR) analysis to analyze the causal effect of the 25-hydroxyvitamin D [25(OH)D] concentration on COVID-19 susceptibility, severity and hospitalization traits by using summary-level GWAS data. The causal associations were estimated with inverse variance weighted (IVW) with fixed effects (IVW-fixed) and random effects (IVW-random), MR-Egger, weighted median and MR Robust Adjusted Profile Score (MR.RAPS) methods. We further applied the MR Steiger filtering method, MR Pleiotropy RESidual Sum and Outlier (MR-PRESSO) global test and PhenoScanner tool to check and remove single nucleotide polymorphisms (SNPs) that were horizontally pleiotropic.

**Results:** We found no evidence to support the causal associations between the serum 25(OH)D concentration and the risk of COVID-19 susceptibility (IVW-fixed: odds ratio [OR] = 0.9049, 95% confidence interval [CI] 0.8197∼0.9988, p = 0.0473), severity (IVW-fixed: OR = 1.0298, 95% CI 0.7699∼1.3775, p = 0.8432) and hospitalized traits (IVW-fixed: OR = 1.0713, 95% CI 0.8819∼1.3013, p = 0.4878) using outlier removed sets at a Bonferroni-corrected p threshold of 0.0167. Sensitivity analyses did not reveal any sign of horizontal pleiotropy.

**Conclusions:** Our MR analysis provided precise evidence that genetically lowered serum 25(OH)D concentrations were not causally associated with COVID-19 susceptibility, severity or hospitalized traits. Our study therefore did not provide evidence assessing the role of vitamin D supplementation during the COVID-19 pandemic. High-quality randomized controlled trials are necessary to explore and define the role of vitamin D supplementation in the prevention and treatment of COVID-19.

## Introduction

The coronavirus disease 2019 (COVID-19) pandemic caused by severe acute respiratory syndrome coronavirus 2 (SARS-CoV-2) has struck globally and is exerting a devastating toll on humans[1]. As of February 12, 2021, this emerging highly infectious disease has spread to six continents at light speed, and there have been 107,252,265 confirmed cases of COVID-19, including 2,355,339 deaths, reported to the World Health Organization (WHO) [2]. The pandemic situation has engulfed the global community into an accelerated search for preventive and therapeutic strategies and has led to calls for widespread vitamin D supplementation. On Dec 17, 2020, the National Institute for Health and Care Excellence (NICE) published an updated rapid review of recent studies on vitamin D and COVID-19 in collaboration with Public Health England and the Scientific Advisory Committee on Nutrition [3]. They support the advice for everyone to take vitamin D supplements to maintain bone and muscle health during the autumn and winter months [3]. The UK government also released and updated new guidance allowing extremely clinically vulnerable people to opt in to receive a free 4-month supply of daily vitamin D supplementation [4].

Humans obtain vitamin D from exposure to sunlight by the action of ultraviolet B on the skin, from their diet, and from dietary supplementation. The vitamin D status is reflected by the level of serum 25-hydroxyvitamin D [25(OH)D], which is produced by the hepatic hydroxylation of vitamin D [5]. The beneficial role of vitamin D in bone growth and maintenance is undisputed and and has influenced practical clinical guidelines and public health policies over the years [6]. However, the evidence supporting the role of vitamin D in other health and disease processes, in particular in acute respiratory tract infection or COVID-19, remains controversial. One meta-analysis of 25 randomized controlled trials (RCTs) of over 11,000 participants showed that vitamin D supplementation could reduce the risk of acute respiratory infections. The protective effects were stronger in those with baseline 25(OH)D concentrations <25 nmol/l [7]. However, another prespecified analysis from the D-Health RCT in more than 20,000 Australian adults recruited from the general population suggested that monthly doses of 60,000 IU of vitamin D did not reduce the risk or severity of acute respiratory tract infections. Although the analysis showed a statistically significant effect on the overall duration of symptoms, the reduction was small (0.5 days) and unlikely to be clinically meaningful [8].

RCTs are the optimal study design to explore and define the role of vitamin D supplementation in preventing the occurrence and severe course of COVID-19, but they have challenges [9]. Mendelian randomization (MR) analysis can overcome the limitations of conventional studies by using single nucleotide polymorphisms (SNPs) as instrumental variables (IVs) for assessing the causal effect of a risk factor (exposure) on an outcome[10]. MR relies on three assumptions: (1) the genetic variant is associated with the exposures; (2) the genetic variant is not associated with confounders; and (3) the genetic variant influences the outcome only through the exposures [11]. A two-sample MR obtains IV-exposure and IV-outcome associations from two different sets of participants. The IVs used in MR are available due to the genome-wide association studies (GWAS) being conducted and high-throughput genomic technologies being developed. Therefore, in this study, we used the MR approach to explore the causal effect of the 25(OH)D concentration on COVID-19 susceptibility, severity and hospitalization traits. This approach can thereby provide estimates of the effect of 25(OH)D while reducing bias due to confounding and reverse causation.

## Methods

We performed a two-sample MR analysis to study the effect of the 25(OH)D concentration on COVID-19 susceptibility, severity and hospitalization traits. Our approach relied upon summary-level GWAS data to obtain MR estimates [11, 12]. We used all SNPs that strongly and independently (R^2^ < 0.001) predicted exposures at genome-wide significance (p < 5E−08). Proxy SNPs (R^2^ > 0.9) from LDlink (https://ldlink.nci.nih.gov/) were used when the SNPs were not available for the outcome [13]. The palindromic SNPs with intermediate allele frequencies (palindromic SNPs referred to the SNPs with A/T or G/C alleles and “intermediate allele frequencies” referred to 0.01<allele frequency<0.30) were excluded from the above selected instrument SNPs. SNPs with a minor allele frequency (MAF) of < 0.01 were also excluded. We also calculated the F statistics for the SNPs to measure the strength of the instruments. IVs with a F statistic less than 10 were excluded and were often labeled “weak instruments” [14]. Further, we used the PhenoScanner tool [15, 16] to check whether any of the selected SNPs were associated with other phenotypes at risk of affecting COVID-19 susceptibility, severity or hospitalization independently of the 25(OH)D concentration. These rigorously selected SNPs were used as the final instrumental SNPs for the subsequent MR analysis.

We retrieved summary data for the association between SNPs and the serum 25(OH)D concentration from the UK Biobank (UKB) with phenotype, genotype and clinical information on 417,580 individuals of European ancestry (age range from 40 to 69 years old) [17]. Individuals were identified by projecting the UKB sample to the first two principal components of the 1,000 Genome Project using Hap Map 3 SNPs with MAF > 0.01 in both data sets. Genotype data were quality-controlled and imputed to the Haplotype Reference Consortium and UK10K reference panels by the UKB group. Genetic variants were extracted with a minor allele count > 5 and imputation score > 0.3 for all individuals and converted genotype probabilities to hard-call genotypes using PLINK2. In total, 8,806,780 variants, including 260,713 SNPs on the X chromosome, were available for analysis. A linear mixed model GWAS was performed to identify the associations between genetic variants and the 25(OH)D concentration. Meta-analysis with SUNLIGHT consortium GWAS results [18] was also performed. Information regarding the quality control and statistical analyses has been reported previously [17].

We obtained estimates of the effect of the 25(OH)D concentration on COVID-19 by obtaining effect coefficients from the above SNPs in GWAS meta-analyses from the COVID-19 Host Genetics Initiative (COVID-19 HGI) [19]. The latest summary statistics were from the fifth round of GWAS meta-analysis shared publicly on January 18,2021. Detailed information on participating studies, quality control, and analyses has been provided on the COVID-19 HGI website (https://www.covid19hg.org/results/). The COVID-19 HGI used different case/control definitions to identify genetic variants associated with COVID-19 susceptibility, disease severity and hospitalized cases of European ancestry and all ancestries. In our study, we only included European ancestry. We used a susceptibility phenotype that compared 38,984 confirmed COVID-19 cases, defined as individuals with laboratory confirmation of SARS-CoV-2 infection based on nucleic acid amplification- or serology-based tests or by electrical health records (using International Classification of Diseases [ICD] or physician notes), chart review or self-reporting, with 1,644,784 controls enrolled in the cohorts and not included as cases. To assess COVID-19 severity, we used the severe phenotype with 5,101 patients, who were defined as patients with COVID-19 with very severe respitatory symptoms and requiring respiratory support, including intubation, CPAP, BiPAP, continuous external negative pressure or high-flow nasal cannula. Controls were 1,383,241 individuals enrolled in the cohorts and not included as cases. The hospitalized phenotype compared 9,986 hospitalized patients with COVID-19, with 1,877,672 controls enrolled in the cohorts and not included as cases. The study details included in the COVID-19 HGI GWAS meta-analyses and three phenotypes are shown in Tables S1-3 in the Supplementary Material. We extracted 114 independent SNPs from the 25(OH)D GWAS as IVs for the three COVID-19 phenotypes. One SNP (rs182244780) was not available in the GWAS of the hospitalized phenotype, and we did not find the proxy SNP from LDlink; therefore, it was left out of the analysis.

After harmonizing the exposure and outcomes datasets, two SNPs (rs11606, rs2246832) were removed for being palindromic with intermediate allele frequencies. Finally, 112, 112 and 111 SNPs were the “Complete sets” involved in the MR analyses (Table 1).

**Table 1.**
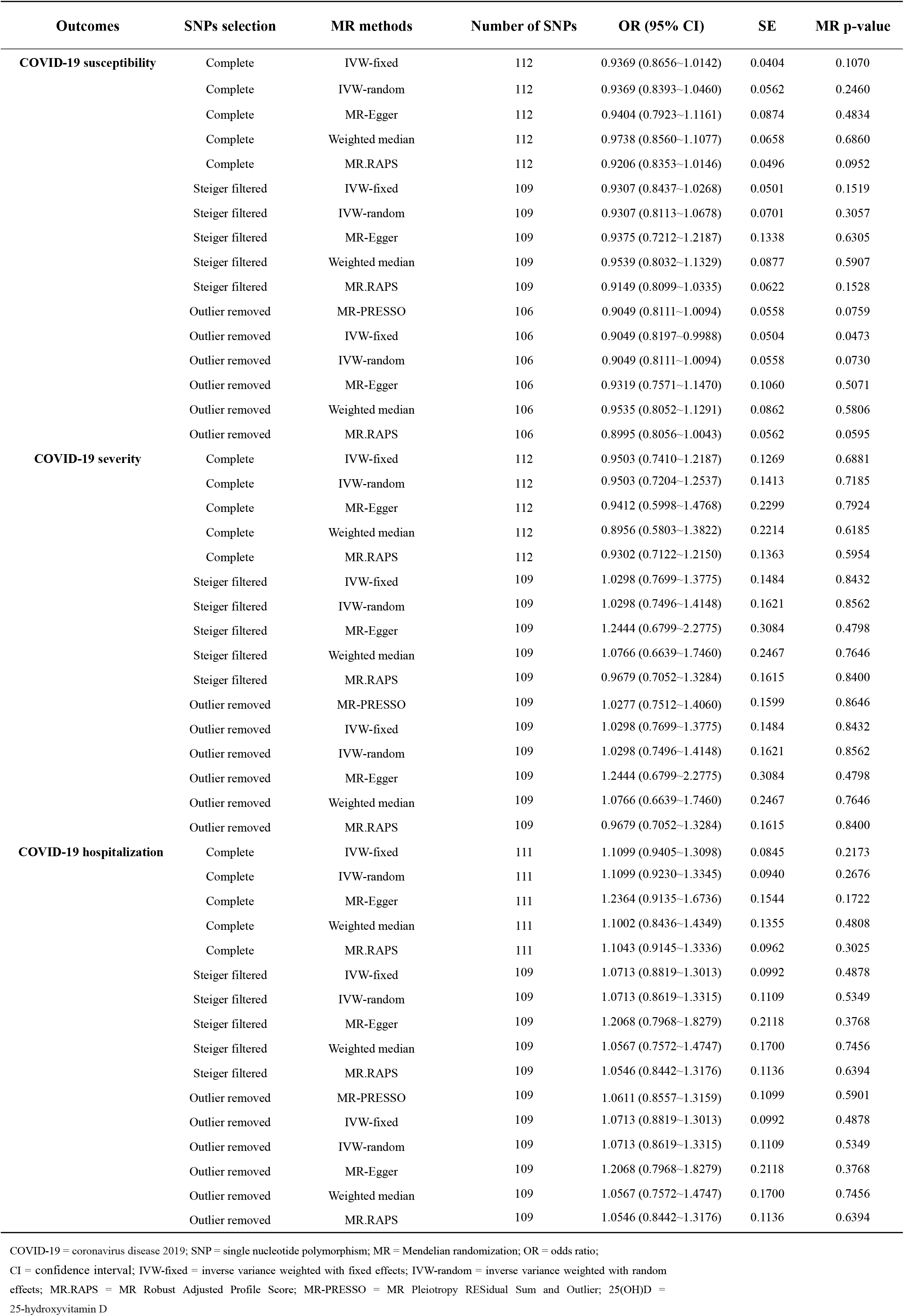
Summary of the univariable Mendelian randomization of the causal effect of the serum 25(OH)D concentration on three COVID-19 phenotypes using different sets of SNPs as instrumental variables.

| Outcomes | SNPs selection | MR methods | Number of SNPs | OR (95% CI) | SE | MR p-value |
| --- | --- | --- | --- | --- | --- | --- |
| COVID-19 susceptibility | Complete | IVW-fixed | 112 | 0.9369 (0.8656~1.0142) | 0.0404 | 0.1070 |
|  | Complete | IVW-random | 112 | 0.9369 (0.8393~1.0460) | 0.0562 | 0.2460 |
|  | Complete | MR-Egger | 112 | 0.9404 (0.7923~1.1161) | 0.0874 | 0.4834 |
|  | Complete | Weighted median | 112 | 0.9738 (0.8560~1.1077) | 0.0658 | 0.6860 |
|  | Complete | MR.RAPS | 112 | 0.9206 (0.8353~1.0146) | 0.0496 | 0.0952 |
|  | Steiger filtered | IVW-fixed | 109 | 0.9307 (0.8437~1.0268) | 0.0501 | 0.1519 |
|  | Steiger filtered | IVW-random | 109 | 0.9307 (0.8113~1.0678) | 0.0701 | 0.3057 |
|  | Steiger filtered | MR-Egger | 109 | 0.9375 (0.7212~1.2187) | 0.1338 | 0.6305 |
|  | Steiger filtered | Weighted median | 109 | 0.9539 (0.8032~1.1329) | 0.0877 | 0.5907 |
|  | Steiger filtered | MR.RAPS | 109 | 0.9149 (0.8099~1.0335) | 0.0622 | 0.1528 |
|  | Outlier removed | MR-PRESSO | 106 | 0.9049 (0.8111~1.0094) | 0.0558 | 0.0759 |
|  | Outlier removed | IVW-fixed | 106 | 0.9049 (0.8197~0.9988) | 0.0504 | 0.0473 |
|  | Outlier removed | IVW-random | 106 | 0.9049 (0.8111~1.0094) | 0.0558 | 0.0730 |
|  | Outlier removed | MR-Egger | 106 | 0.9319 (0.7571~1.1470) | 0.1060 | 0.5071 |
|  | Outlier removed | Weighted median | 106 | 0.9535 (0.8052~1.1291) | 0.0862 | 0.5806 |
|  | Outlier removed | MR.RAPS | 106 | 0.8995 (0.8056~1.0043) | 0.0562 | 0.0595 |
| COVID-19 severity | Complete | IVW-fixed | 112 | 0.9503 (0.7410~1.2187) | 0.1269 | 0.6881 |
|  | Complete | IVW-random | 112 | 0.9503 (0.7204~1.2537) | 0.1413 | 0.7185 |
|  | Complete | MR-Egger | 112 | 0.9412 (0.5998~1.4768) | 0.2299 | 0.7924 |
|  | Complete | Weighted median | 112 | 0.8956 (0.5803~1.3822) | 0.2214 | 0.6185 |
|  | Complete | MR.RAPS | 112 | 0.9302 (0.7122~1.2150) | 0.1363 | 0.5954 |
|  | Steiger filtered | IVW-fixed | 109 | 1.0298 (0.7699~1.3775) | 0.1484 | 0.8432 |
|  | Steiger filtered | IVW-random | 109 | 1.0298 (0.7496~1.4148) | 0.1621 | 0.8562 |
|  | Steiger filtered | MR-Egger | 109 | 1.2444 (0.6799~2.2775) | 0.3084 | 0.4798 |
|  | Steiger filtered | Weighted median | 109 | 1.0766 (0.6639~1.7460) | 0.2467 | 0.7646 |
|  | Steiger filtered | MR.RAPS | 109 | 0.9679 (0.7052~1.3284) | 0.1615 | 0.8400 |
|  | Outlier removed | MR-PRESSO | 109 | 1.0277 (0.7512~1.4060) | 0.1599 | 0.8646 |
|  | Outlier removed | IVW-fixed | 109 | 1.0298 (0.7699~1.3775) | 0.1484 | 0.8432 |
|  | Outlier removed | IVW-random | 109 | 1.0298 (0.7496~1.4148) | 0.1621 | 0.8562 |
|  | Outlier removed | MR-Egger | 109 | 1.2444 (0.6799~2.2775) | 0.3084 | 0.4798 |
|  | Outlier removed | Weighted median | 109 | 1.0766 (0.6639~1.7460) | 0.2467 | 0.7646 |
|  | Outlier removed | MR.RAPS | 109 | 0.9679 (0.7052~1.3284) | 0.1615 | 0.8400 |
| COVID-19 hospitalization | Complete | IVW-fixed | 111 | 1.1099 (0.9405~1.3098) | 0.0845 | 0.2173 |
|  | Complete | IVW-random | 111 | 1.1099 (0.9230~1.3345) | 0.0940 | 0.2676 |
|  | Complete | MR-Egger | 111 | 1.2364 (0.9135~1.6736) | 0.1544 | 0.1722 |
|  | Complete | Weighted median | 111 | 1.1002 (0.8436~1.4349) | 0.1355 | 0.4808 |
|  | Complete | MR.RAPS | 111 | 1.1043 (0.9145~1.3336) | 0.0962 | 0.3025 |
|  | Steiger filtered | IVW-fixed | 109 | 1.0713 (0.8819~1.3013) | 0.0992 | 0.4878 |
|  | Steiger filtered | IVW-random | 109 | 1.0713 (0.8619~1.3315) | 0.1109 | 0.5349 |
|  | Steiger filtered | MR-Egger | 109 | 1.2068 (0.7968~1.8279) | 0.2118 | 0.3768 |
|  | Steiger filtered | Weighted median | 109 | 1.0567 (0.7572~1.4747) | 0.1700 | 0.7456 |
|  | Steiger filtered | MR.RAPS | 109 | 1.0546 (0.8442~1.3176) | 0.1136 | 0.6394 |
|  | Outlier removed | MR-PRESSO | 109 | 1.0611 (0.8557~1.3159) | 0.1099 | 0.5901 |
|  | Outlier removed | IVW-fixed | 109 | 1.0713 (0.8819~1.3013) | 0.0992 | 0.4878 |
|  | Outlier removed | IVW-random | 109 | 1.0713 (0.8619~1.3315) | 0.1109 | 0.5349 |
|  | Outlier removed | MR-Egger | 109 | 1.2068 (0.7968~1.8279) | 0.2118 | 0.3768 |
|  | Outlier removed | Weighted median | 109 | 1.0567 (0.7572~1.4747) | 0.1700 | 0.7456 |
|  | Outlier removed | MR.RAPS | 109 | 1.0546 (0.8442~1.3176) | 0.1136 | 0.6394 |

COVID-19 = coronavirus disease 2019; SNP = single nucleotide polymorphism; MR = Mendelian randomization; OR = odds ratio; CI = confidence interval; IVW-fixed = inverse variance weighted with fixed effects; IVW-random = inverse variance weighted with random effects; MR.RAPS = MR Robust Adjusted Profile Score; MR-PRESSO = MR Pleiotropy RESidual Sum and Outlier; 25(OH)D = 25-hydroxyvitamin D

We applied the principles of univariable two-sample MR to obtain estimates of the causal effect of the 25(OH)D concentration on COVID-19 susceptibility, severity and hospitalization traits separately. Analyses were performed using the TwoSampleMR R package of MR-Base[12] (https://github.com/MRCIEU/TwoSampleMR). The statistical tests of the MR analysis were two-sided, and the results of the MR analyses were considered statistically significant at a Bonferroni-corrected p*<* 0.0167 (e.g., 0.05/3 outcomes). The causal associations were estimated with inverse variance weighted (IVW) with fixed effects (IVW-fixed) and random effects (IVW-random) [12, 20], MR-Egger [21, 22] and Weighted median estimate methods [23]. The IVW method uses a meta-analysis approach to combine the Wald ratios of the causal effects of each SNP and can provide the most precise estimates [12, 20]. The Weighted median estimate provides a reliable effect estimate of the causal effect when at least 50% of the weight in the analysis comes from effective IVs [23]. MR-Egger regression is used to create a weighted linear regression of the outcome coefficients with the exposure coefficients. The slope of the weighted regression line provides an asymptotically unbiased causal estimate of the exposure on the outcome if the INSIDE (instrument strength is independent of direct effect) assumption is met. In addition, the intercept of the MR-Egger regression line was used to quantify the amount of horizontal pleiotropy present in the data averaged across the genetic instruments [21, 22]. Under the INSIDE assumption, the MR-Egger intercept test identifies horizontal pleiotropy if the intercept from the MR-Egger analysis is not equal to zero [22]. We also calculated the Robust Adjusted Profile Score (MR.RAPS) to estimate the causal effects, which can lead to considerably higher statistical power than the conventional MR analysis [24]. MR.RAPS considers the measurement error in SNP-exposure effects and is unbiased when there are many weak instruments as well as robust to systematic and idiosyncratic pleiotropy [24]. The MR.RAPS method can also alleviate but cannot solve the problem of horizontal pleiotropy [24].

When selecting SNPs from the GWAS, especially in very large GWAS, it can be difficult to determine whether a SNP has its primary association with the exposure being studied or the outcome [25]. For example, in our study, if COVID-19 phenotypes exert a causal effect on the serum 25(OH)D concentration, then there is a possibility that some SNPs primarily associated with COVID-19 may also pass the genome-wide significance threshold in a GWAS of the 25(OH)D concentration with a large sample size. These SNPs can then misleadingly be applied as genetic instruments for the 25(OH)D concentration when they should actually be applied as IVs for COVID-19. Therefore, we applied MR Steiger filtering [26] as implemented in the TwoSampleMR R package to test the causal direction of each of the extracted SNPs on the exposures [25(OH)D] and outcome (COVID-19 phenotypes). This approach calculates the variance explained in the exposure and the outcome by the instrumenting SNPs and tests if the variance in the outcome is less than the exposure. For any SNP that had less variance in the exposure than the outcome (which means it showed evidence of primarily affecting outcomes rather than exposures), we removed those lipid SNPs and conducted IVW, MR-Egger, Weighted median and MR.RAPS using the remaining instruments (“Steiger filtered” sets). IVs with “TRUE” meant the evidence for causality in the expected direction, while “FALSE” meant the evidence for causality in the reverse direction (Tables S4-6). After Steiger filtering, 109, 109 and 109 SNPs (rs182244780, rs1894100 and rs4694423 were excluded) were involved in the MR analysis (Table 1).

We further applied the MR Pleiotropy RESidual Sum and Outlier (MR-PRESSO) global test [27] to reduce heterogeneity in the estimate of the causal effect to remove SNPs that were horizontal pleiotropic outliers. We conducted this analysis by using the MR-PRESSO R package (https://github.com/rondolab/MR-PRESSO). The number of distributions was set to 1,000 and the threshold was set to 0.05. We also conducted IVW, MR-Egger, Weighted median and MR.RAPS using the remaining instruments (“Outlier removed” sets). None of the horizontal pleiotropic outliers were identified for the COVID-19 severity and hospitalization, while three SNPs (rs532436, rs12949853 and rs72997688) were removed for COVID-19 susceptibility (Table 1). We estimated the power of our study using the outlier removed sets according to a method suggested by Brion et al [28]. The equations using an approximate linear model on the observed binary (0-1) scale were adapted for binary outcomes, which require several parameters to estimate. These parameters include the proportion of phenotypic variation explained by IV SNPs, the effect size of the exposure to the outcome at the epidemiological level, sample size, and the standard deviation (SD) of the exposure and outcome. The three outlier removed sets of SNPs for COVID-19 susceptibility, severity and hospitalization collectively explained 0.0183, 0.0185 and 0.0185 of the variance in the 25(OH)D concentration, respectively (Tables S4-6).

We used the IVW, MR-Egger and Maximum likelihood [29] methods to detect heterogeneity. Heterogeneity was quantified by the Cochran Q statistic. To identify potentially influential SNPs, we performed a “leave-one-out” sensitivity analysis in which we excluded one SNP at a time and performed an IVW-random method on the remaining SNPs to identify the potential influence of outlying variants on the estimates. To validate the MR results, we conducted sensitivity analyses using genetic variants associated with 25(OH)D that were identified in the SUNLIGHT consortium GWAS results [18]. The large, multicenter, GWAS analysis considering phenotype data from 79,366 individuals of European ancestry included 31 studies from epidemiological cohorts from Europe, Canada, and the USA [18]. We first extracted 7 SNPs associated with 25(OH)D at genome-wide significance (p < 5E−08) and pruned for linkage disequilibrium at a R^2^ coefficient of correlation of < 0.001 (Table S7). Then, to reduce the limitations of SNP numbers, we pruned SNPs to a R^2^ of 0.01 and extracted 10 SNPs associated with 25(OH)D (Table S8). We performed IVW-fixed, IVW-random [12, 20], MR-Egger [21, 22] and Weighted median methods [23] to calculate the causal estimates.

Further, we used the PhenoScanner tool [15, 16] to check whether any of the 109 identified SNPs in the outlier removed sets in the current study were associated with other phenotypes at risk of affecting the three COVID-19 phenotypes independent of the 25(OH)D concentration. We assessed SNPs at a threshold of p < 5E-08 for their association with any other phenotypes. Using the PhenoScanner tool, we found that 35 SNPs (rs10454087, rs1047891, rs10822145, rs12056768, rs12317268, rs1260326, rs142004400, rs1800588, rs182050989, rs2037511, rs2131925, rs2229742, rs2418929, rs2642439, rs2756119, rs28407950, rs2952289, rs3745669, rs41301394, rs429358, rs4418728, rs4616820, rs4846917, rs512083, rs532436, rs5771043, rs58387006, rs6834488, rs71297391, rs73413596, rs804281, rs8063565, rs8107974, rs964184 and rs9861009) were significantly associated with hematological traits (e.g., white blood cell count, platelet count and granulocyte count). Changes in hematological parameters in SARS-CoV-2 infected patients are very common, and several studies have shown that hematological parameters are markers of disease severity and progression. It was confirmed that SARS-CoV-2 infection was associated with alterations in the blood cell count. One in four COVID-19 patients experienced some form of leukopenia (WBC < 4.00E+09 cells/l), with the majority (63.0%) exhibiting lymphocytopenia (lymphocyte count < 1.00E+09 cells/l) [30]. Additionally, a reduced number of eosinophils has been reported in more than half (52.9%) of patients who tested positive for COVID-19 [31]. Zhou F et al [32] also reported a significant platelet count reduction between patients with severe disease and those exhibiting mild disease. To avoid horizontal pleiotropy caused by possible causal mechanistic associations between hematological traits and COVID-19 phenotypes, we performed univariable two-sample MR with IVW-fixed and IVW-random methods to assess the epidemiological correlations between hematological traits and COVID-19 phenotypes. Genome-wide significant (p < 5E−08) and independent (R^2^ < 0.001) genetic variants of predicted exposures of 34 blood cell indices, including white blood cell count, platelet count and lymphocyte count were extracted from the Astle et al. GWAS [33]. The GWAS on blood cell traits was performed on a population of approximately 173,000 individuals of European ancestry, excluding those with blood cancers or major blood disorders, largely from the UK. The significance threshold was set using Bonferroni correction at p < 4.90E-04 (0.05/[34×3]). We excluded SNPs significantly associated with blood cell traits if causal links were identified between blood cell traits and COVID-19 phenotypes.

## Results

As we mentioned above, outlier removed sets included 106,109 and 109 SNPs, respectively (Table 1). The detailed characteristics of SNPs in outlier removal sets associated with the 25(OH)D concentration are shown in Tables S4-6. For these IVs, all of the F statistics were above 10 (ranging from 29.7798 to 1349.6840 for COVID-19 susceptibility; ranging from 29.7798 to 1349.6840 for COVID-19 severity; and ranging from 29.7798 to 1349.6840 for COVID-19 hospitalization) with average F statistics of 89.1985, 87.7882 and 87.7882, respectively (Tables S4-6), indicating that they satisfy the strong relevance assumption of MR and that weak instrument bias would not substantially influence the estimations of causal effects. The results of MR analyses of the 25(OH)D concentration and three COVID-19 phenotypes with three sets of SNPs (complete, Steiger filtered and outlier removed sets) are presented in Table 1. We estimated the overall odds ratio (OR) of COVID-19 phenotypes per 1-SD increase in the 25(OH)D concentration. IVW-fixed, IVW-random, MR-Egger, Weighted median and MR.RAPS using the complete sets of SNPs did not suggest the causal effect of increased/decreased 25(OH)D concentration on COVID-19 susceptibility, severity or hospitalization traits at a Bonferroni-corrected p threshold of 0.0167 (IVW-fixed: OR = 0.9369, 95% confidence interval [CI] 0.8656∼1.0142, p = 0.1070; MR-Egger: OR = 0.9404, 95% CI 0.7923∼1.1161, p = 0.4834; Weighted median: OR = 0.9738, 95% CI 0.8560∼1.1077, p = 0.6860; MR.RAPS: OR = 0.9206, 95% CI 0.8353∼1.0146, p = 0.0952 for COVID-19 susceptibility) (Table 1). The results did not provide evidence for the causal effects, and were stable and consistent using the Steiger filtered (IVW-fixed: OR = 0.9307, 95% CI 0.8437∼1.0268, p = 0.1519 for COVID-19 susceptibility) and outlier removed sets (IVW-fixed: OR = 0.9049, 95% CI 0.8197∼0.9988, p = 0.0473 for COVID-19 susceptibility) for the three COVID-19 traits (Table 1 and Figure 1).

**Figure 1.**
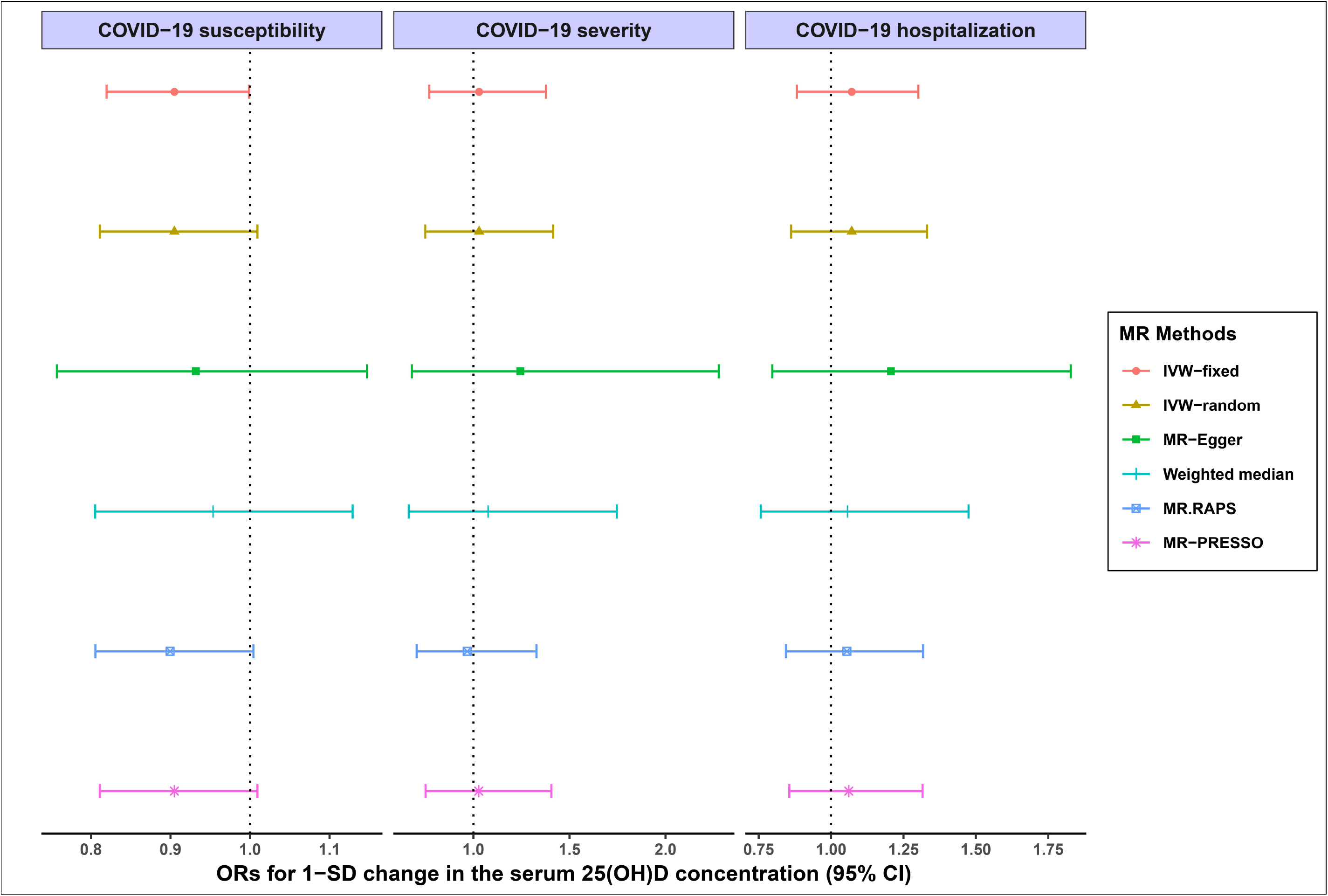
Forest plot of the causal effects of the 25(OH)D concentration on COVID-19 susceptibility, severity and hospitalization traits. The analysis was conducted using outlier removed sets of SNPs. OR = odds ratio; SD = standard deviation; CI = confidence interval; IVW-fixed = inverse variance weighted with fixed effects; IVW-random = inverse variance weighted with random effects; MR.RAPS = MR Robust Adjusted Profile Score; MR-PRESSO = MR Pleiotropy RESidual Sum and Outlier; COVID-19 = coronavirus disease 2019; 25(OH)D = 25-hydroxyvitamin

Table 2 displays the heterogeneity analysis results using the IVW, MR-Egger and Maximum likelihood methods, and pleiotropy analysis using the MR-Egger intercept test. At the Bonferroni-corrected p threshold of 0.0167, we observed strong evidence of heterogeneity across complete SNP sets for COVID-19 susceptibility (IVW: Q = 214.2548, p = 1.49E-08, MR-Egger: Q = 214.2489, p = 1.06E-08, Maximum likelihood: Q = 214.2201, p = 1.50E-08), but evidence of heterogeneity was lacking for the COVID-19 severity and hospitalization (IVW: Q = 137.6576, p = 0.0439, MR-Egger: Q = 137.6540, p = 0.0382, Maximum likelihood: Q = 137.6563, p = 0.0439 for COVID-19 severity; IVW: Q = 136.2421, p = 0.0456, MR-Egger: Q = 135.2766, p = 0.0447, Maximum likelihood: Q = 136.2287, p = 0.0456 for COVID-19 hospitalization). After Steiger filtering and outliers removing, we did not observe the evidence of heterogeneity using three SNP sets for the three COVID-19 phenotypes (for COVID-19 susceptibility using outlier removed sets, IVW: Q = 128.5356, p = 0.0592, MR-Egger: Q = 128.4032, p = 0.0525, Maximum likelihood: Q = 128.4887, p = 0.0595). Based on the MR-Egger intercept test, we did not find evidence of directional pleiotropy between the 25(OH)D concentration and three COVID-19 phenotypes using three SNPs sets (complete SNPs sets: intercept = −0.0001, p = 0.9561 for COVID-19 susceptibility; intercept = 0.0033, p = 0.9573 for COVID-19 severity; intercept = −0.0037, p = 0.3797 for COVID-19 hospitalization).

**Table 2.** Heterogeneity and pleiotropy analysis of the serum 25(OH)D concentration on three COVID-19 phenotypes using different sets of SNPs as instrumental variables.

| Outcomes | SNPs selection | MR methods | Number of SNPs | Cochran Q statistic | Heterogeneity p-value | MR-Egger |  |
| --- | --- | --- | --- | --- | --- | --- | --- |
|  |  |  |  |  |  | Intercept | Intercept p-value |
| COVID-19 susceptibility | Complete | IVW | 112 | 214.2548 | 1.49E-08 | -0.0001 | 0.9561 |
|  | Complete | MR-Egger | 112 | 214.2489 | 1.06E-08 |  |  |
|  | Complete | Maximum likelihood | 112 | 214.2201 | 1.50E-08 |  |  |
|  | Steiger filtered | IVW | 109 | 211.3969 | 1.08E-08 | -0.0002 | 0.9495 |
|  | Steiger filtered | MR-Egger | 109 | 211.3890 | 7.64E-09 |  |  |
|  | Steiger filtered | Maximum likelihood | 109 | 211.3606 | 1.09E-08 |  |  |
|  | Outlier removed | IVW | 106 | 128.5356 | 0.0592 | -0.0008 | 0.7440 |
|  | Outlier removed | MR-Egger | 106 | 128.4032 | 0.0525 |  |  |
|  | Outlier removed | Maximum likelihood | 106 | 128.4887 | 0.0595 |  |  |
| COVID-19 severity | Complete | IVW | 112 | 137.6576 | 0.0439 | 0.0033 | 0.9573 |
|  | Complete | MR-Egger | 112 | 137.6540 | 0.0382 |  |  |
|  | Complete | Maximum likelihood | 112 | 137.6563 | 0.0439 |  |  |
|  | Steiger filtered | IVW | 109 | 128.7771 | 0.0843 | -0.0053 | 0.4718 |
|  | Steiger filtered | MR-Egger | 109 | 128.1526 | 0.0780 |  |  |
|  | Steiger filtered | Maximum likelihood | 109 | 128.7767 | 0.0843 |  |  |
|  | Outlier removed | IVW | 109 | 128.7771 | 0.0843 | -0.0053 | 0.4718 |
|  | Outlier removed | MR-Egger | 109 | 128.1526 | 0.0780 |  |  |
|  | Outlier removed | Maximum likelihood | 109 | 128.7767 | 0.0843 |  |  |
| COVID-19 hospitalization | Complete | IVW | 111 | 136.2421 | 0.0456 | -0.0037 | 0.3797 |
|  | Complete | MR-Egger | 111 | 135.2766 | 0.0447 |  |  |
|  | Complete | Maximum likelihood | 111 | 136.2287 | 0.0456 |  |  |
|  | Steiger filtered | IVW | 109 | 134.9943 | 0.0403 | -0.0033 | 0.5101 |
|  | Steiger filtered | MR-Egger | 109 | 134.4456 | 0.0375 |  |  |
|  | Steiger filtered | Maximum likelihood | 109 | 134.9891 | 0.0403 |  |  |
|  | Outlier removed | IVW | 109 | 134.9943 | 0.0403 | -0.0033 | 0.5101 |
|  | Outlier removed | MR-Egger | 109 | 134.4456 | 0.0375 |  |  |
|  | Outlier removed | Maximum likelihood | 109 | 134.9891 | 0.0403 |  |  |

COVID-19 = coronavirus disease 2019; SNP = single nucleotide polymorphism; MR = Mendelian randomization; OR = odds ratio; IVW = inverse variance weighted; 25(OH)D = 25-hydroxyvitamin D

We performed “leave-one-out” analysis based on the IVW-random method using the outlier removed sets for three COVID-19 phenotypes and found that there was no potentially influential SNP driving the causal link, and the result was stable (Tables S9-11). Extended MR analysis using 25(OH)D-related variants from the SUNLIGHT consortium GWAS [18] also reported no evidence of causal association between 25(OH)D and COVID-19 traits (Tables S7-8). To avoid horizontal pleiotropy, we performed univariable two-sample MR between hematological traits and COVID-19 phenotypes due to 35 SNPs associated with blood traits. Using the IVW-fixed and IVW-random methods, we found no evidence to report the significant causal associations between 34 hematological traits and three COVID-19 phenotypes (p<4.90E-04) (e.g., white blood cell count with COVID-19 susceptibility, beta = 0.0016, IVW-fixed p = 0.0650, IVW-random p = 0.0629) (Tables S12-14). Therefore, we did not remove those 35 SNPs.

Table 3 shows the sample size in the current analysis for the three COVID-19 traits. The total COVID-19 susceptibility group sample size was 1,683,768, of which 38,984 were COVID-19 cases. The overall COVID-19 severity group sample size was 1,388,342, of which 5,101 were COVID-19 severe cases. The sample size of all COVID-19 hospitalization was 1,887,658, of which 9,986 were COVID-19 hospitalized cases. We presented the power estimations for a range of proportions of 25(OH)D variation explained by genetic variants. Under the current sample size with outlier removed sets, our study had 80% power to detect a causal effect of a relative 12% (ORs less than 0.8994) decrease in COVID-19 susceptibility risk, a relative 32% (ORs less than 0.7251) decrease in COVID-19 severity risk and a relative 22% (ORs less than 0.8004) decrease in COVID-19 hospitalization risk per 1-SD increase in the 25(OH)D concentration.

**Table 3.** Number of COVID-19 cases and controls and statistical power in a Mendelian randomization study of the serum vitamin D concentration and three COVID-19 phenotypes.

| Exposure | Outcome |  |  |  |  | Minimum detectable odds ratio |  |  |  |  |
| --- | --- | --- | --- | --- | --- | --- | --- | --- | --- | --- |
|  | Trait | Cases | Controls | Total | Proportion of cases | r<sup>2</sup> = 0.01 | r<sup>2</sup> = 0.02 | r<sup>2</sup> = 0.03 | r<sup>2</sup> = 0.04 | r<sup>2</sup> = 0.05 |
| 25(OH)D | COVID-19 susceptibility | 38984 | 1644784 | 1683768 | 0.0232 | 1.1430/0.8579 | 1.1010/0.8994 | 1.0834/0.9178 | 1.0722/0.9288 | 1.0639/0.9356 |
|  | COVID-19 severity | 5101 | 1383241 | 1388342 | 0.0037 | 1.3945/0.6115 | 1.2789/0.7251 | 1.2249/0.7754 | 1.1971/0.8031 | 1.1742/0.8239 |
|  | COVID-19 hospitalization | 9986 | 1877672 | 1887658 | 0.0053 | 1.2795/0.7179 | 1.1976/0.8004 | 1.1613/0.8390 | 1.1397/0.8588 | 1.1249/0.8737 |

COVID-19 = coronavirus disease 2019; 25(OH)D = 25-hydroxyvitamin D; r<sup>2</sup>= the proportion of phenotypic variation explained by genetic variants Minimum detectable odds ratio per 1-SD increase/decrease in 25(OH)D concentration: assume 80% power, 5% alpha level and that 1-5% of 25(OH)D variance is explained by the genetic variants used in the current study.

## Discussion

There is no doubt that immune modulation is the fulcrum of most diseases, and COVID-19 is no exception. Vitamin D has many mechanisms to modulate immunity, such as physical barriers and innate and adaptive immunity, to reduce the risk of microbial and viral infection [34, 35]. Vitamin D was reported to help maintain tight junctions, gap junctions, and adherens junctions to resist infections [36]. It can enhance cellular innate immunity through the release of antimicrobial peptides, such as human cathelicidin [37], which exhibits direct antimicrobial activities by killing invading pathogens by perturbing the cell membranes and viral envelope, such as SARS-CoV-2 [38]. Vitamin D could also activate T cells, B cells, dendritic cells and macrophages by the vitamin D receptor (VDR) expressed by most immune cells [35]. These cells are able to produce a wide repertoire of responses that ultimately determine the nature and duration of the immune responses. Moreover, vitamin D exerts an inhibitory, anti-inflammatory action on the adaptive immune system. Vitamin D can reduce the production of proinflammatory T helper type 1 (Th1) cytokines, such as tumor necrosis factor α and interferon γ, and increase the expression of anti-inflammatory cytokines by macrophages [39]. It can also promote cytokine production by T helper type 2 (Th2) cells, which helps enhance the suppression of Th1 cells indirectly [40].

In this MR study, we used the strong IVs from the summary statistics of the largest GWAS conducted for vitamin D and COVID-19 phenotypes in European populations. We aimed to determine whether the relationship between 25(OH)D and COVID-19 was causal by employing a range of two-sample MR methods. We employed a range of methods known to control for pleiotropy, checked the heterogeneity and obtained highly consistent results. Pleiotropic effects were detected by using the MR-Egger intercept and MR-PRESSO method. Using the MR design, we could mitigate the confounding factors due to the application of Mendel’s second law of the random assortment of alleles. Reverse causality was also prevented because genetic variants were fixed at conception and cannot be affected by disease processes. We used the PhenoScanner tool to detect potential pleiotropic SNPs. Some SNPs were found to be associated with hematological parameters. We then performed univariable MR and found no evidence to support the causal association between hematological parameters and COVID-19 traits. The results above showed that the presence of pleiotropic SNPs was minimal. Taken together, our MR results did not support the evidence that the serum 25(OH)D concentration was a causal factor for COVID-19 susceptibility, severity or hospitalization traits. Our study therefore did not provide evidence assessing the role of vitamin D supplementation during the pandemic.

This is the first study to illustrate the causal relationship between the serum 25(OH)D concentration and COVID-19 using MR analysis. Multiple published traditional epidemiology studies have demonstrated the role of vitamin D supplementation in reducing the risk of COVID-19 and its severity. However, the results were inconsistent and likely to be confounded by multiple unmeasured or improperly controlled variables. Hastie et al [41, 42] used individual data from UK Biobank participants to correlate historical vitamin D levels checked between 2006 and 2010 with risk for COVID-19 positivity. They found that the 25(OH)D concentration was univariably associated with severe COVID-19 and mortality, but statistical significance was lost after adjustment for confounders. A retrospective, observational analysis including 191,779 patients from across all US states used data from tests performed at a national clinical laboratory [43]. They found a strong association between lower 25(OH)D concentrations and an increased rate of SARS-CoV-2 positivity. This remained significant after adjustment for sex, age, latitude and ethnicity. A retrospective cohort from Chicago included 489 patients tested for COVID-19 who had a vitamin D level checked in the year before testing. The relative risk of testing positive for COVID-19 was 1.77 times greater for patients with likely deficient vitamin D status (25-hydroxycholecalciferol < 20 ng/ml) compared with patients with likely sufficient vitamin D status, a difference that was statistically significant [44]. Carpagnano et al [45] analyzed the demographic, clinical and laboratory data of 42 patients with acute respiratory failure due to COVID-19 in Italy. They reported that patients with severe vitamin D deficiency had a significantly higher mortality risk. However, the small sample size and low follow-up of enrolled patients might limit the power of the study. Despite major efforts to control for confounding, such observational studies still remained prone to residual confounding by uncontrolled or imperfectly measured covariates. Additionally, collider bias could arise when researchers restrict analyses on a collider variable in observational studies. The collider bias referred to restricting analyses to some people who experienced a specific event, such as hospitalization with COVID-19, or volunteered their participation in a large-scale study. The association effects in the specific sample will not be a reliable indication of the individual-level causal effects. Therefore, collider bias caused associations to fail to generalize beyond the sample and for causal inferences to be inaccurate within the sample [46]. Reverse causal effects would also violate the results of observational studies. Vitamin D binding protein was reported as a positive acute phase reactant after infections, which might lower the level of 25(OH)D. A decrease in 25(OH)D may also be the result of COVID-19 [47].

In one quasi-experimental study, Annweiler et al [48] included sixty-six residents with COVID-19 from a French nursing home. They reported that vitamin D supplementation represented an effective, accessible and well-tolerated treatment for COVID-19 to reduce the severe COVID-19 cases and improve survival rates. To the best of our knowledge, most RCTs of vitamin D in the community are unlikely to be complete until spring 2021 [49], although we noted the positive results of a randomized trial[50] reporting that the administration of a high dose of calcifediol or 25(OH)D significantly reduced the need for intensive care unit (ICU) treatment of patients requiring hospitalization due to COVID-19. However, imperfect double-blinding, uneven distribution and imperfect control for potential confounders were limitations of this trial. Well-designed studies are paramount for more fully exploring and defining the role of vitamin D supplementation in preventing the occurrence and severe course of COVID-19.

Despite the MR design being less susceptible to confounding than other observational studies, limitations still exist. First, potential pleiotropy is the common limitation on all MRs, and our results may still have been affected by unmeasured horizontal pleiotropy. To assess this bias, we assessed potential pleiotropy using the MR-Egger method and MR-PRESSO method. We also used MR Steiger filtering and the PhenoScanner tool and observed no evidence that pleiotropic SNPs existed. Hence, whereas the risk of a residual horizontal pleiotropic effect cannot be ruled out, we still believe it is unlikely to change the conclusions of this study in a clinically meaningful way. Second, the low power of MR might be the reason for null results rather than a true null. However, our study had 80% power to detect the decreased risk of COVID-19 susceptibility, severity and hospitalization traits with ORs of 0.89, 0.72 and 0.80 per SD increase in the serum 25(OH)D concentration. Third, genetic associations may be affected by population stratification, which is another potential bias-inducing factor for MR analyses since the MAF differences among different ancestries and may differ between the populations in the exposure and outcome GWAS. The limitation was minimized by utilizing genetic associations from studies mainly of people of European ancestry with genomic control in our study. However, extending our results to people outside Europe should proceed with caution. Lastly, the MR study only tested the linear effect of serum vitamin D concentration in the general population. Future studies may be designed to comprehensively evaluate any nonlinear relationships between the vitamin D concentration and COVID-19 traits.

## Conclusion

In summary, we provided precise evidence that genetically lowered serum 25(OH)D concentrations were not causally associated with COVID-19 susceptibility, severity or hospitalization traits. In balance, the current evidence does not support the need to take the vitamin D supplements in order to reduce the risk of COVID-19 susceptibility, severity and hospitalization. We are looking forward to the results of ongoing well-designed randomized controlled trials providing more evidence concerning the role of vitamin D in the prevention and treatment of COVID-19.

## Data Availability

All datasets presented in this study are included in the article/ supplementary material.

https://www.example.com

## Acknowledgement

The authors wish to thank Dr Siying Zhuang for the support in the preparation of this manuscript.

## Funding statement

This work was supported by grants from Beijing Municipal Science and Technology Commission

(Z181100001718195).

## Authors’ contributions

Yun Tian and Zhiyong Cui conceptualized and designed the study. Zhiyong Cui provided the package codes in R language and analyzed the data in the study. Zhiyong Cui drafted the manuscript. Yun Tian gave constructive suggestions when writing the manuscript. Both two authors have read the manuscript.

